# Machine Learning Prediction of Disease Trajectories for Children with Juvenile Idiopathic Arthritis

**DOI:** 10.64898/2026.04.18.26351165

**Authors:** Seungwon Lee, Marie Davidian, Marc D. Natter, Bryce B. Reeve, Laura E. Schanberg, Eleanor Belkin, Min-Lee Chang, Yukiko Kimura, Mei-Sing Ong

## Abstract

**Background:** Despite advances in therapy, optimal management of juvenile idiopathic arthritis (JIA) remains challenging. The ability to predict disease progression in JIA can improve personalized treatment decisions, but few reliable clinical predictors have been identified. We developed machine learning approaches to predict disease trajectories in children with JIA.

**Methods:** Using data from the Childhood Arthritis and Rheumatology Research Alliance (CARRA) Registry (years 2015-2024), we developed machine learning models to predict attainment of inactive disease in children with non-systemic JIA. We applied Dynamic Bayesian Networks (DBN) to model temporal dependencies and causal relationships, and Convolutional Neural Networks (CNN) to capture complex non-linear patterns. Model input included demographic factors, longitudinal clinical factors, and medication use in the preceding 12 months.

**Findings:** A total of 8,093 participants were included. When tested on an independent test cohort, both DBN (AUC:0.76; precision:0.73; recall:0.83; F1-score:0.78; accuracy:0.71) and CNN (AUC:0.76; precision:0.71; recall:0.63; F1-score:0.67; accuracy:0.70) models achieved comparable performance in predicting inactive disease. Disease activity levels in the preceding 12 months, presence of enthesitis and uveitis were the strongest predictors. Causal relationships captured in the DBN model revealed suboptimal care patterns, likely shaped by insurance constraints and a predominantly reactive approach to JIA management.

**Interpretation:** Our study demonstrates that machine learning approaches can predict disease trajectories in JIA with good discriminative performance. Unlike prior studies that predict outcomes at single timepoints, our models are the first to predict inactive disease longitudinally. However, suboptimal care patterns in retrospective data limit models’ capacity to learn treatment-outcome relationships, underscoring critical opportunities to improve JIA care and the need for prospective comparative studies to better inform prediction models.

**Funding:** Patient-Centered Outcomes Research Institute (PCORI) Award (ME-2022C2-25573-IC).

**RESEARCH IN CONTEXT:** 

**Evidence before this study:** Numerous studies have sought to identify clinical predictors of JIA progression and outcomes. However, few reliable predictors have emerged and existing prediction models demonstrate limited performance. As a result, our ability to personalize treatment decisions based on individual risk of severe disease course remains limited.

**Added value of this study:** We developed novel machine learning models that predict individualized disease trajectories in children with polyarticular and oligoarticular JIA using data from their preceding 12-month clinical course. These models demonstrated strong discriminative performance and outperformed previously published machine learning approaches in JIA. Unlike prior studies limited to single time-point predictions, our models are the first to predict inactive disease longitudinally, enabling a patient-specific projection of disease progression over time. Importantly, our findings also bright to light patterns of suboptimal care, likely driven by insurance constraints and a reactive treatment paradigm, underscoring critical opportunities to improve JIA management.

**Implications of all the available evidence:** Our models have the potential to support clinical decision-making by enabling early identification of children with JIA at risk for unfavorable disease trajectories. In addition, the suboptimal care patterns and systems-level barriers identified through our analyses highlight priority areas for quality improvement initiatives and policy interventions to reduce gaps in JIA care delivery.

## INTRODUCTION

Despite advances in therapy, optimal management of juvenile idiopathic arthritis (JIA) remains challenging. Published evidence suggests a “therapeutic window of opportunity”, wherein early intervention increases the likelihood of clinical remission.^1–3^ However, treatment benefits must be balanced against its risks. Although biologic treatments have significantly improved outcomes, they expose patients to potential adverse effects, including serious infections and malignancies.^4^ The ability to predict an individual’s likelihood of following a more severe JIA trajectory could improve personalized treatment decision-making. However, although several studies have examined clinical predictors of JIA outcomes, few reliable predictors have been identified.^5^

In this study, we applied machine learning approaches to predict disease trajectories in children with polyarticular and oligoarticular JIA, with the goal of providing personalized risk stratification to inform therapeutic decision-making. Beyond prediction alone, we sought to identify longitudinal clinical factors influencing disease progression and providing insights into drivers of adverse outcomes. We applied two complementary machine learning methods: Dynamic Bayesian Networks (DBN) for modeling temporal dependencies and potential causal relationships, and Convolutional Neural Networks (CNN) for capturing complex non-linear patterns in longitudinal data.

## METHODS

### Study participants and data source

We extracted data from the Childhood Arthritis and Rheumatology Research Alliance (CARRA) Registry (ClinicalTrials.gov Identifier: NCT02418442) – the largest prospective safety and research registry of childhood rheumatic diseases in North America with 74 participating academic medical centers in the United States and Canada.^6^ Study participants comprised children with JIA who were enrolled in the CARRA Registry from 2015-2024, excluding those with systemic JIA. Clinical data were collected from participants approximately every 6 months at routine clinic visits. We excluded participants who did not have disease activity data entered between 12 and 36 months from enrollment. Data captured by the registry include demographics, diagnoses, longitudinal disease activity, comorbidities, health-related quality of life and other patient-reported outcomes, medications with start/stop dates, adverse events, and laboratory results.

This study was approved by the Boston Children’s Hospital Institutional Review Board (IRB-P00030533).

### Study Outcome

The primary outcome of interest was the prediction of attainment of inactive disease status over time. Disease activity was defined by clinical Juvenile Arthritis Disease Activity Score (cJADAS10), a three-variable composite score for JIA that includes active joint count (maximum 10 joints), physician global assessment of disease activity, and parent/child global assessment of well-being.^7^ cJADAS10 scores range from 0 to 30, with higher values indicating greater disease activity. Published cJADAS10 cutoffs define levels of disease activity in polyarticular JIA (≤2.5 inactive; >2.5-5 minimal; >5-16 moderate; and >16 high disease activity) and oligoarticular JIA (≤1.1 inactive disease; >1.1-4 minimal; >4-12 moderate; and >12 high disease activity.^8^

### Analysis

We applied two machine learning approaches to predict disease trajectories over time – Dynamic Bayesian Networks (DBN) and Convolutional Neural Network (CNN).

DBNs are probabilistic graphical models that extend traditional Bayesian Networks to capture temporal relationships in sequential data.^9^ In a DBN, variables at each time point are represented as nodes, with directed edges between nodes indicating probabilistic dependencies between the variables (Supplementary Figure 1). For example, a directed edge from nodes A to B indicates that A “influences” B; in other words, the presence of A alters the probability of B occurring. These dependencies can exist both within a time point and across successive time periods. For disease trajectory prediction, DBNs learn transition probabilities that govern how clinical states evolve over time, providing an interpretable causal architecture of disease progression that can be validated against medical knowledge.

In contrast, CNN is a specialized class of artificial neural networks. that mimic aspects of visual processing in the human brain, using layers that detect increasingly complex features as information flows through the system.^10^ The core building blocks include convolutional layers that apply mathematical filters to detect regional patterns in data. Multiple convolutional layers with different filter sizes can capture patterns at various temporal scales, from short-term fluctuations to long-term trends.^11–13^ While CNNs operate as “black boxes” with limited interpretability, they excel at modelling complex, non-linear relationships that often pose challenges for DBNs. As such, they serve as a strong performance benchmark when compared to state-of-the-art deep learning methods.

### Model Development

#### Dynamic Bayesian Network

In-depth description of the approach for developing DBN is presented in Supplementary Methods. We systematically evaluated candidate clinical variables for model inclusion, first applying a data quality threshold by excluding variables that were incomplete for more than 50% of visits across all participants. From the remaining variables, we selected those demonstrating meaningful temporal relationships with disease activity through network connectivity analysis, excluding variables that formed isolated nodes without significant connections to other clinical measures. Continuous variables were discretized into clinically meaningful categories, as defined in consultation with clinical stakeholders. Missing values were handled by assigning them to a separate “Unknown” category.

In addition to prediction of disease activity, we applied the derived DBN to identify factors leading to attainment of inactive disease. The graphical component of DBN provides a visual tool for identifying temporal dependencies among variables, including direct and indirect relations, and their evolution over time. Furthermore, the topology of the graph describes Markovian properties of variables that allow us to decompose the network into related modules. The local Markov property asserts that a node is independent of its non-descendant nodes, given the parent nodes.^14^ Thus, for a given variable of interest, we can identify a minimal set of other variables (neighboring nodes) that influence, or are affected by, the variable. This minimal set, known as the “Markov blanket”, represents distinct patterns of interacting factors contributing to the outcome of interest (Supplementary Figure 1).

#### Convolutional Neural Network (CNN)

We investigated multiple CNN architectures to identify the optimal model configuration. Our exploration included: (1) a standard feedforward neural network, (2) a fully convolutional network that eliminates fully connected layers, and (3) a dual-branch architecture with separate input pathways. Details of these modelling approaches are described in Supplementary Methods.

We first developed CNN models using the full set of variables (see Supplementary Table 1). To mitigate potential overfitting, we subsequently developed models using a subset of features identified to be predictive by the DBN models. In all analyses, continuous variables were log-transformed to reduce skewness, and categorical variables were encoded using one-hot encoding. Supplementary Table 3 summarizes the hyperparameters used in model development. To further mitigate overfitting, we also adopted two regularization techniques: L1 regularization and dropout.^15,16^

### Model input

We extracted 41 variables from the CARRA Registry, deemed to be important for predicting JIA disease outcomes, based on input from clinical and patient stakeholders (Supplementary Table 1). Longitudinal clinical data are recorded in the registry at six-month intervals, and detailed start and stop dates for all medications are available. We categorized medications taken by participants into three major groups: biologic/targeted synthetic disease-modifying antirheumatic (b/tsDMARD), conventional synthetic DMARD (csDMARD), and systemic glucocorticoids. Supplementary Table 2 summarizes the list of medications in each category. Medication use was represented as a binary variable, with exposure marked as true if it occurred during the specified period.

To identify the optimal temporal window for modeling, we systematically evaluated various look-back periods (i.e. the span of historical data used as input), ranging from 6 to 36 months prior to the prediction point. Our analysis revealed that remission probability is predominantly influenced by disease activity patterns within the preceding 12 months, with earlier clinical history more than 12 months ago providing minimal additional predictive value. Consequently, we adopted a 12-month lookback window that captures clinically relevant temporal dependencies while optimizing model complexity and computational performance.

Given that the participants had registry visits approximately every 6 months, inputs to the model therefore included clinical variables at T-6 and T-12 months before the point of prediction. Additionally, we included patient demographics as static variables and medication data from two periods: 6 to 12 months before prediction (T-12 to T-6) and the immediate 6 months before prediction (T-6 to T). The inclusion of these 2 windows enables us to examine the following question: How do preceding patterns of treatment affect subsequent disease activity?

#### Model performance and validation

We randomly partitioned the dataset into training (80%) and test sets (20%) to enable unbiased assessment of model generalizability.^17^ Model development and validation were conducted using 5-fold cross-validation on the training set, with final evaluation performed on the independent test set. For all analyses, we used AUC (Area Under the Receiver Operating Characteristic) as the main performance measure for model comparison, and additionally evaluated precision, recall, F1-score and accuracy.

## RESULTS

Of 12,064 participants with JIA captured in the CARRA Registry, 8,093 participants satisfied the inclusion criteria and were included in our analyses (Table 1). The mean and median visits per participant between 12 and 36 months were 4 and 3 visits, respectively.

**Table 1.** Study cohort (n=8,093)

| Attribute | N (%) |
| --- | --- |
| Sex |  |
| Female | 5837 (72.1) |
| Male | 2256 (27.9) |
| Age |  |
| ≤ 6 | 1843 (22.8) |
| >6 to ≤12 | 2821 (34.9) |
| >12 | 3429 (42.4) |
| JIA Category |  |
| Enthesitis-related arthritis | 900 (11.1) |
| Oligoarthritis | 3169 (39.2) |
| Polyarthritis (RF –ve) | 2609 (32.2) |
| Polyarthritis (RF +ve) | 573 (7.1) |
| Psoriatic arthritis | 614 (7.6) |
| Undifferentiated arthritis | 228 (2.8) |

### Dynamic Bayesian Network

Of 41 candidate variables extracted from the CARRA Registry (Supplementary Table 1), 26 variables either had more than 50% missing data or consistently exhibited zero network connectivity (i.e. no connections to other variables in the network), and were therefore excluded. The final set of variables used for the development of DBN included the following: (a) demographics (age, sex); (b) clinical attributes (JIA category, anti-CCP positivity, HLAB-27 positivity, family history of rheumatic disease, time from symptom onset to diagnosis, cJADAS10, uveitis, active enthesitis, active sacroiliitis, radiographic joint damage); and (c) medications (bDMARD, csDMARD, systemic glucocorticoids).

Figure 1 depicts the derived DBN model using the full dataset. The model predicted attainment of disease remission at time T with an average AUC of 0.76 (precision: 0.73; recall: 0.83; F1-score: 0.78; accuracy: 0.71), given clinical data at T-6 and T-12, and medication data in the preceding 12 months. The AUC of the model when predicting disease remission at specific time points, between 12 and 36 months from registry enrollment, ranged from 0.74 (at 12 months) to 0.77 (at 24 and 30 months) (Figure 2). The average AUC ranged from 0.74 to 0.79 across different JIA categories (Table 2).

**Figure 1.**
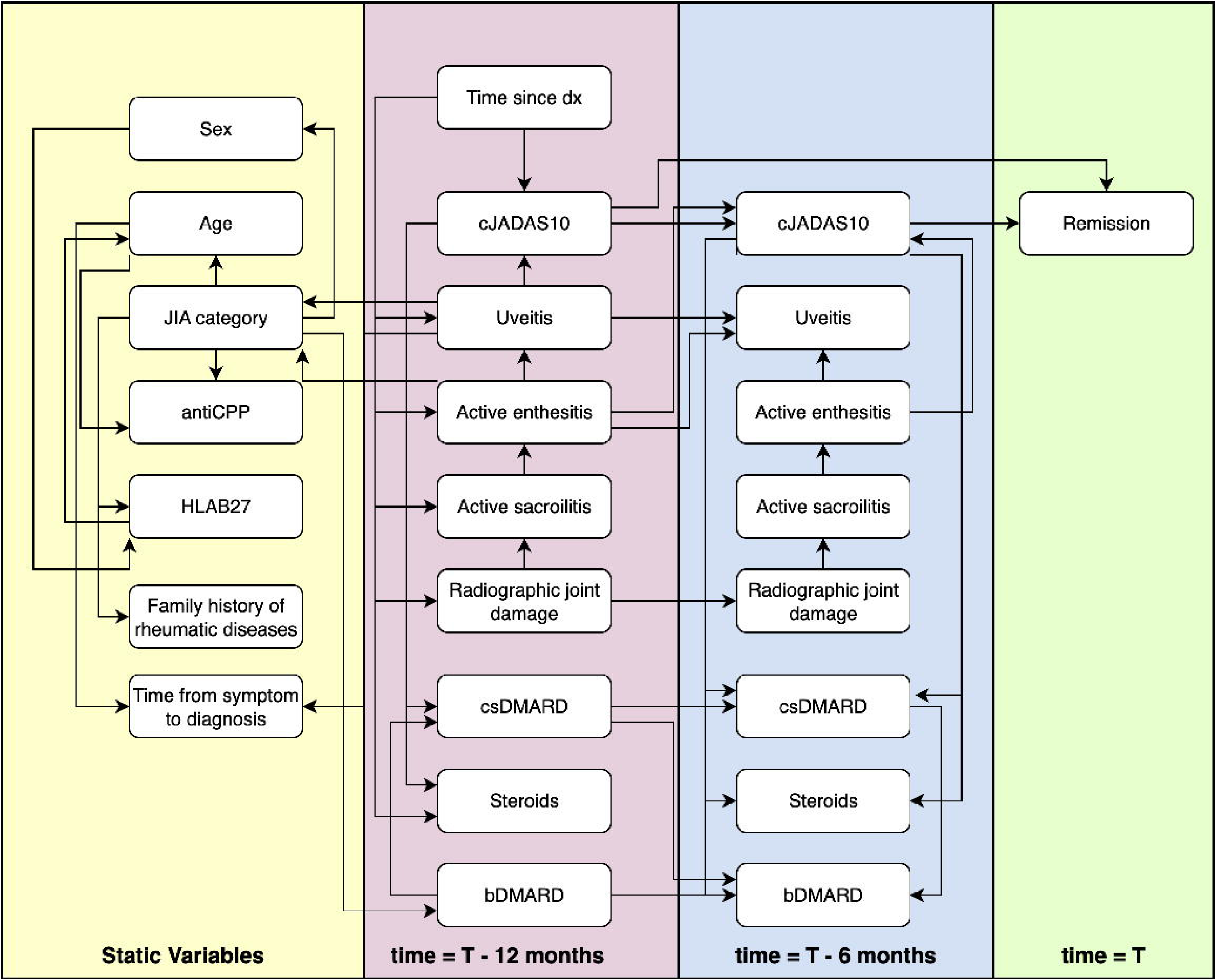
DBN model for predicting cJADAS10 remission at time T.

**Figure 2.**
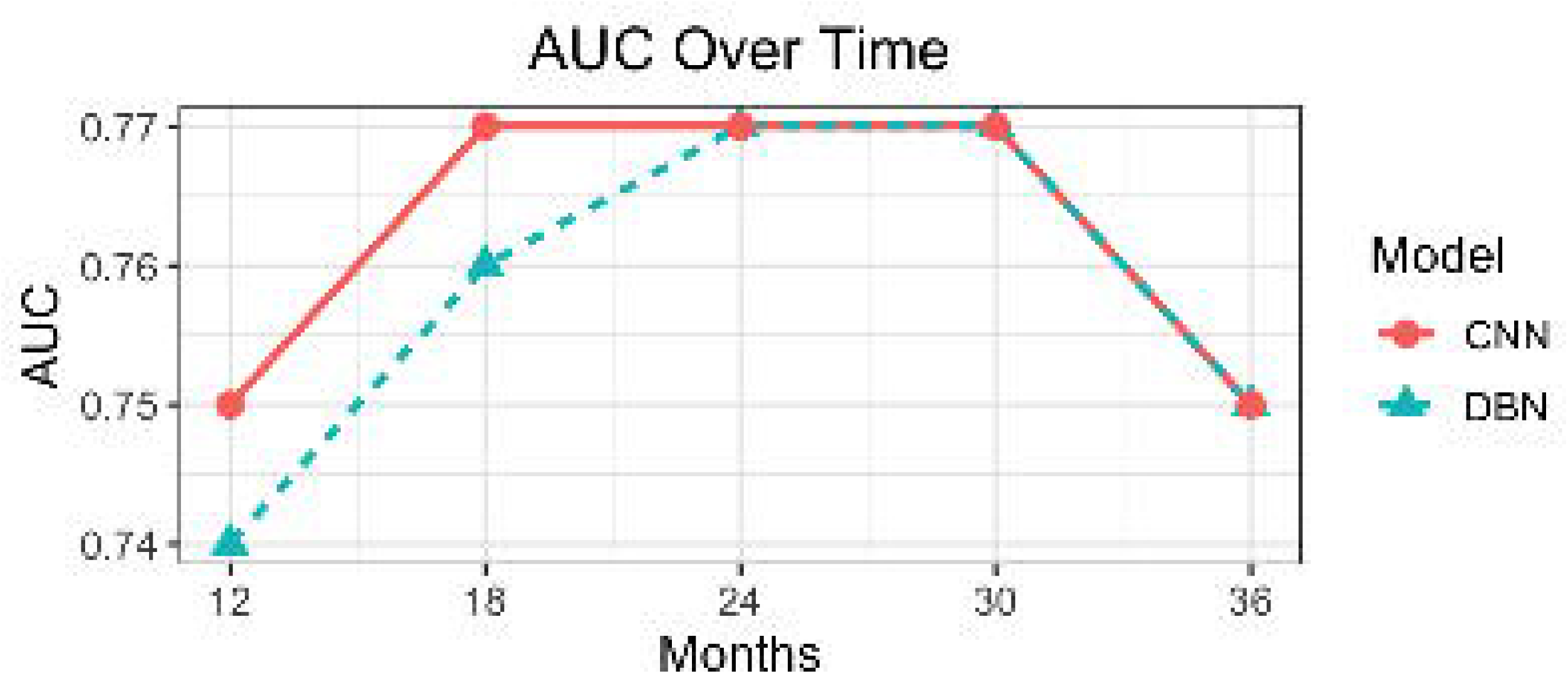
Predictive performance of DBN and CNN models over time.

**Table 2.** Average AUC of models for predicting remission by JIA categories.

| JIA category | DBN | CNN |
| --- | --- | --- |
| Enthesitis-related arthritis | 0.78 | 0.78 |
| Oligoarthritis | 0.74 | 0.71 |
| Polyarthritis (RF negative) | 0.75 | 0.76 |
| Polyarthritis (RF positive) | 0.75 | 0.74 |
| Psoriatic arthritis | 0.76 | 0.75 |
| Undifferentiated arthritis | 0.79 | 0.75 |

Disease activity at T-6 and T-12 months predicted future remission and were the only variables included in the Markov blanket ^14^ of remission at time T, indicating their direct influence on remission outcomes. Analysis of the Markov blanket of disease activity at T-12 months showed that clinical variables influencing increased disease activity at T-12 included active enthesitis and new-onset uveitis at T-12.

Disease activity, in turn, influenced the treatment received by a participant in the subsequent follow-up period. Analysis of the Markov blanket of disease activity level at T-6 months showed similar patterns – level of disease activity at T-6 months was influenced by disease activity at T-12 months and the presence of active enthesitis in a subset of patients. Again, we observed that disease activity influenced the treatment received by a participant in the subsequent follow-up period. Treatment patterns were also influenced by treatment patterns in the prior time slice. We saw that past levels of disease activity directly influenced the probability of remission at time T. In both time periods, prior treatment patterns did not appear to influence disease activity. Of note, we further observed that the initiation of bDMARD was influenced by patients’ JIA category, rather than by the level of disease activity.

### Convolutional Neural Network

All three CNN architectures exhibited comparable performance, with AUC ranging from 0.76 to 0.77 (Supplementary Table 3). Therefore, we chose to use fully convolutional neural networks for the primary analysis of disease trajectory prediction, given their design simplicity. Because the kernel of a convolutional layer processes information from local regions of the input data before passing it to the next layer, a temporal convolutional network that incorporates all input variables is mathematically equivalent to a single-branch feedforward neural network replicated across all time points.

Training AUC rapidly approached 1.0 within the first few epochs, indicating the models were learning to perfectly classify training data. However, validation and test AUC peaked at 0.75-0.77 in early training (typically epochs 3-5), then declined and plateaued around 0.70 for the remainder of training (Supplementary Figure 2). This substantial divergence between training and validation performance indicates severe overfitting. L1 regularization and dropout did not improve model performance or reduce the training-validation gap (Supplementary Figure 2).

Given the persistent overfitting despite regularization, we hypothesized that the high dimensionality of our feature set relative to our sample size was the primary cause. To address this, we employed DBN as a data-driven feature selection method to identify the most predictive variables. We then retrained the fully CNN model using only the DBN-selected features. With this feature-reduced approach, training and validation AUC converged at approximately 0.76, with minimal divergence throughout training. As illustrated in Figure 3, the final CNN model demonstrated performance comparable to that of the DBN model, with an average AUC of 0.76 (precision: 0.71; recall: 0.63; F1-score: 0.67; accuracy: 0.70). The average AUC ranged from 0.71 to 0.78 across different JIA categories (Table 2).

### Sensitivity analyses

We conducted multiple sensitivity analyses to assess model robustness and identify potential improvements (see Supplementary Methods). However, none of these analyses improved the performance of either the DBN or CNN models beyond the baseline AUC of 0.76. Notably, analyses that reduced the training sample size consistently resulted in worse performance. This included experiments with stricter cohort selection criteria (which excluded certain patient subgroups) and models trained to predict outcomes at single time points rather than multiple time points (resulting in fewer training examples per model). Sensitivity analyses exploring alternative feature representations (e.g. encoding medication use as cumulative exposure versus binary indicators) did not alter model performance. Imputing missing data using autoencoder, MICE MissForest, and MIDAS resulted in decreased performance (measured by AUC and accuracy) for both DBN and CNN models. In the final models, missing data were handled by assigning them to a distinct category.

## DISCUSSION

We have developed machine learning models capable of predicting disease trajectories of individual children with polyarticular and oligoarticular JIA, based on their preceding 12-month clinical course. Both DBN and CNN models achieved comparable performance, attaining an AUC of 0.76, indicating acceptable to good discriminative ability. Given the heterogeneity of JIA phenotypes, the complex nature of disease progression, and the absence of reliable biomarkers, this level of performance is clinically meaningful and has the potential to support clinical decision-making alongside physician and caregiver judgment.

Our models outperformed most published machine learning studies in JIA^18–20^ and performed comparably to one study on the Nordic cohort,^21^ though variability in outcome definitions across studies limits direct comparison. We chose cJADAS10 as the outcome of interest because it represents a comprehensive, validated assessment of JIA disease activity recommended by international consensus groups and widely adopted in clinical trials,^22,23^ making our findings broadly applicable. The Nordic model applied logistic regression with 7 variables (erythrocyte sedimentation rate [ESR], c-reactive protein [CRP], morning stiffness, physician global assessment, ANA, HLA-B27, and ankle joint arthritis) to predict non-achievement of remission off medications and achieved an AUC of 0.78 (IQR 0.72-0.82). However, we were unable to replicate these results in our dataset, achieving an AUC of 0.71 (precision: 0.53; recall: 0.13; F1-score: 0.21; accuracy: 0.76) using the same approach and variables (excluding ankle joint arthritis, which is not captured in the CARRA Registry) to predict cJADAS10. Importantly, while the goal of previous studies was to predict outcomes at isolated timepoints based on baseline clinical predictors, our models are the first to predict disease trajectories.

Beyond trajectory prediction, the graphical structure of the DBN model provides insights into factors driving disease remission over time. Specifically, prior disease activity levels emerged as the strongest predictors of remission. This is consistent with a prior study reporting that physician’s global assessment and count of active joints – both components of cJADAS10 – were the most important features for predicting inactive disease at 24 months, thus highlighting the importance of regular assessment of disease activity in the monitoring of disease course.^20^ Other clinical variables influencing increased disease activity included active enthesitis and new-onset uveitis. These observations are consistent with current clinical understanding. The presence of enthesitis (or belonging to the enthesitis-related arthritis category) is associated with a more severe or persistent disease course, compared with other JIA categories.^24,25^ Uveitis has also been shown to correlate with higher treatment burden and disease activity,^26^.

The causal relationships captured by the DBN model also suggests that the observed treatment patterns in our cohort resulted from clinicians responding to disease activity, rather than treatment directly determining disease outcomes, mirroring the reactive nature of treatment decision-making in real-world JIA management whereby therapies are escalated in response to signs of clinical worsening, rather than proactively to prevent future flares. This approach means that treatment typically lags behind disease activity, which may result in suboptimal outcomes. Prediction tools that anticipate disease progression before clinical deterioration occurs have the potential to support earlier, more proactive treatment decisions that may improve long-term outcomes. However, the predominantly reactive treatment patterns reflected in the current data may limit the ability of predictive models to learn optimal treatment-outcome relationships. This limitation highlights both the challenges inherent in developing robust prediction models from retrospective data and the critical need for prospective comparative studies to better inform prediction models.

We further observed that b/tsDMARD initiation was driven primarily by JIA category rather than disease activity level. Clinicians are likely to initiate b/tsDMARD treatment earlier in patients with JIA categories associated with poorer prognoses, such as RF-positive polyarticular JIA. However, this pattern could also stem from insurance policies that establish category-specific treatment pathways.^27^ Insurance prior authorization requirements also vary by JIA category, with more restrictive criteria for expensive b/tsDMARDs in perceived milder categories such as oligoarticular JIA, compared to aggressive categories such as polyarticular RF-positive disease. Insurers also routinely deny biologic therapy for sacroiliitis and ERA in patients under age 16, which may have contributed to worse outcomes in these patients. Additionally, most insurance policies require patients to demonstrate inadequate response to csDMARDs before approving b/tsDMARDs. This stepwise treatment pattern was reflected in the DBN model, where prior csDMARD use influenced subsequent initiation of b/tsDMARD. Such mandated delays may lead to missed opportunities for early intervention during critical windows when disease-modifying treatment could be most effective.^3,28^

Our analyses, therefore, highlight clinical practice and systems-level barriers to optimal treatment of JIA. The comparable performance of 2 distinct modelling approaches suggests that these constraints may impose a performance ceiling that limits any predictive approach. Due to the reactive approach to managing JIA, current practice patterns captured in real-world data may not represent the most optimal treatment strategies. Furthermore, when treatment decisions are primarily determined by step therapy mandates and coverage restrictions rather than clinical considerations, algorithms cannot learn true treatment-outcome relationships, because such relationships are not adequately represented in the training data.

The similar performance of DBN and CNN models may also stem from the inherent challenges of predicting outcomes for an uncommon disease such as JIA. Our dataset captures many variables, but was drawn from a relatively small patient cohort. This data structure typically favors DBNs. In contrast, CNNs typically require larger datasets to effectively learn complex temporal patterns. As shown in our analyses, CNN models frequently achieved near-perfect performance on the training data, but performed poorly on the test sets – an indication of overfitting. However, test set performance improved significantly when the number of input variables was reduced, using features identified through DBN.

Our study has several limitations. First, substantial missing data affected key variables shown to be predictive in prior studies, including ESR, CRP, and morning stiffness. Second, our dataset did not capture several clinically important variables identified in other studies, such as specific joints involved and symmetric joint distribution. Inclusion of these variables has the potential to improve model performance. A technical limitation is that the structure learning algorithm in DBN models prioritizes strong predictive relationships, potentially excluding variables with modest effect sizes or those providing redundant information. Consequently, some clinically relevant but weaker predictors may not be represented in the final model structure. This trade-off favors interpretability but may not capture the full complexity of disease progression. Conversion of continuous variables into categorical variables may also reduce the predictive accuracy of the model.

Nonetheless, our study has important strengths. We developed the first machine learning models to predict disease trajectories in JIA, demonstrating clinically meaningful performance despite the complexity and heterogeneity of the disease. Beyond prediction, our DBN model provided interpretable insights into temporal factors influencing remission, while also revealing how clinical practice and healthcare system barriers may constrain optimal treatment delivery, thus limiting the ability to model treatment-outcome relationships. Prospective investigations that are now underway will provide additional information (e.g. the “STep-up and step-down therapeutic strategies in childhood ARthritiS (STARS) trial (NCT03728478), a study designed to compare the effectiveness of conventional, treat-to-target approach, with that of an early aggressive intervention). The results of this and similar studies are expected to generate high-quality evidence to inform treatment decisions and will also provide robust data for the development of future prediction models.

## Data Availability

De-identified participant data from the CARRA Registry that support the findings of this study are available from the Childhood Arthritis and Rheumatology Research Alliance (CARRA) subject to the CARRA Data and Sample Sharing Policy. Data access requires submission of a data request and approval by the CARRA Data, Sample, and Publications Committee. Investigators may apply through the CARRA Research Portal according to the procedures outlined in the CARRA data and sample sharing policy.

## Acknowledgement

Data were obtained from the Childhood Arthritis and Rheumatology Research Alliance (CARRA) Registry. The authors acknowledge the contributions of the CARRA Registry investigators, research coordinators, and participating patients and families. A full list of CARRA site investigators is available at carragroup.org. The analyses, conclusions, and opinions expressed herein are solely those of the authors and do not necessarily reflect those of CARRA.

## REFERENCES

1 Wallace CA, Giannini EH, Spalding SJ, et al. Clinically inactive disease in a cohort of children with new-onset polyarticular juvenile idiopathic arthritis treated with early aggressive therapy: time to achievement, total duration, and predictors. J Rheumatol 2014; 41: 1163–70.

2 Ong M-S, Ringold S, Kimura Y, Schanberg L, Tomlinson G, Natter M. Improved disease course associated with early initiation of biologics in untreated polyarticular Juvenile Idiopathic Arthritis: A trajectory analysis of the STOP-JIA study. Arthritis Rheumatol 2021.

3 Ringold S, Ong M-S, Tomlinson G, et al. Three-Year Outcomes and Latent Class Trajectory Analysis of the Childhood Arthritis and Rheumatology Research Alliance Polyarticular JIA Consensus Treatment Plans Study. Arthritis Rheumatol 2025; 77: 1433–41.

4 Hashkes PJ, Uziel Y, Laxer RM. The safety profile of biologic therapies for juvenile idiopathic arthritis. Nat Rev Rheumatol 2010; 6: 561–71.

5 Shoop -Worrall SJW, Wu Q, Davies R, Hyrich KL, Wedderburn LR. Predicting disease outcomes in juvenile idiopathic arthritis: challenges, evidence, and new directions. Lancet Child Adolesc Health 2019; 3: 725–33.

6 Fuhlbrigge RC, Schanberg LE, Kimura Y. CARRA: the childhood arthritis and rheumatology research alliance. Rheum Dis Clin North Am 2021; 47: 531–43.

7 Consolaro A, Giancane G, Schiappapietra B, et al. Clinical outcome measures in juvenile idiopathic arthritis. Pediatr Rheumatol Online J 2016; 14: 23.

8. Trincianti C, van Dijkhuize EHP, Alongi A, et al. Definition and validation of the American College of Rheumatology 2021 JADAS cutoffs for Disease Activity States in Juvenile Idiopathic Arthritis. Arthritis Rheumatol In press 2021.

9 Koller D, Sahami M. Toward optimal features selection. Proceedings of the International Conference on Machine Learning 1996; : 284–292.

10 Derry A, Krzywinski M, Altman N. Convolutional neural networks. Nat Methods 2023; 20: 1269–70.

11 Pelletier C, Webb G, Petitjean F. Temporal convolutional neural network for the classification of satellite image time series. Remote Sens (Basel) 2019; 11: 523.

12 Wibawa AP, Utama ABP, Elmunsyah H, Pujianto U, Dwiyanto FA, Hernandez L. Time-series analysis with smoothed Convolutional Neural Network. J Big Data 2022; 9: 44.

13 Lea C, Vidal R, Reiter A, Hager GD. Temporal convolutional networks: A unified approach to action segmentation. In: Hua G, Jégou H, eds. Computer vision – ECCV 2016 workshops. Cham: Springer International Publishing, 2016: 47–54.

14 Koller D, Friedman N. Probabilistic Graphical Models: Principles And Techniques (adaptive Computation And Machine Learning), 1st edn. Cambridge, MA: The Mit Press, 2009.

15 Moradi R, Berangi R, Minaei B. A survey of regularization strategies for deep models. Artif Intell Rev 2020; 53: 3947–86.

16 Srivastava N, Hinton G, Krizhevsky A. Dropout: a simple way to prevent neural networks from overfitting. The journal of machine 2014.

17 Russell SJ (Stuart J, Norvig Peter, Davis Ernest. Artificial intelligence: A modern approach, 3rd edn. Upper Saddle River, 2009.

18 Guzman J, Henrey A, Loughin T, et al. Predicting Which Children with Juvenile Idiopathic Arthritis Will Not Attain Early Remission with Conventional Treatment: Results from the ReACCh-Out Cohort. J Rheumatol 2019; 46: 628–35.

19 van Dijkhuizen EHP, Aidonopoulos O, Ter Haar NM, et al. Prediction of inactive disease in juvenile idiopathic arthritis: a multicentre observational cohort study. Rheumatology (Oxford) 2018; 57: 1752–60.

20 Reboll o-Giménez AI, Ridella F, Orsi SM, et al. Forecasting Achievement of Inactive Disease in Juvenile Idiopathic Arthritis with Artificial Intelligence. Children (Basel) 2025; 12. DOI:10.3390/children12060741.

21 Rypdal V, Arnstad ED, Aalto K, et al. Predicting unfavorable long-term outcome in juvenile idiopathic arthritis: results from the Nordic cohort study. Arthritis Res Ther 2018; 20: 91.

22 Ringold S, Angeles-Han ST, Beukelman T, et al. 2019 American College of Rheumatology/Arthritis Foundation Guideline for the Treatment of Juvenile Idiopathic Arthritis: Therapeutic Approaches for Non-Systemic Polyarthritis, Sacroiliitis, and Enthesitis. Arthritis Rheumatol 2019; 71: 846–63.

23 Ravelli A, Consolaro A, Horneff G, et al. Treating juvenile idiopathic arthritis to target: recommendations of an international task force. Ann Rheum Dis 2018; 77: 819–28.

24 Taxter AJ, Wileyto EP, Behrens EM, Weiss PF. Patient-reported Outcomes across Categories of Juvenile Idiopathic Arthritis. J Rheumatol 2015; 42: 1914–21.

25 Balay -Dustrude E, Weiss NS, Sutton A, Shenoi S. Predictors of disease activity in patients with juvenile idiopathic arthritis at 12 and 24 months after diagnosis. ACR Open Rheumatol 2024; 6: 489–96.

26 Rypdal V, Glerup M, Songstad NT, et al. Uveitis in Juvenile Idiopathic Arthritis: 18-Year Outcome in the Population-based Nordic Cohort Study. Ophthalmology 2021; 128: 598–608.

27 Shenoi S, Horneff G, Aggarwal A, Ravelli A. Treatment of non-systemic juvenile idiopathic arthritis. Nat Rev Rheumatol 2024; 20: 170–81.

28 Ong MS, Ringold S, Kimura Y, et al. Improved disease course associated with early initiation of biologics in polyarticular juvenile idiopathic arthritis: trajectory analysis of a childhood arthritis and rheumatology research alliance consensus treatment plans study. Arthritis Rheumatol 2021; 73: 1910–20.

