## Supplementary Figure 1 for "Machine Learning Prediction of Disease Trajectories for Children with Juvenile Idiopathic Arthritis"

**Supplementary Figure 1. Bayesian Network**

Diagram A illustrates a simple Bayesian network. Each node corresponds to an attribute of interest, and influence of one node on another is depicted as a directed edge. Given a target variable, we can identify a minimal set of neighboring nodes that influence, or are affected by, the variable. This minimal set, known as the “Markov blanket”, includes the parent, children, and co-parent nodes. In the example below, the Markov blanket of D = {B, E, F}. Diagram B illustrates a Dynamic Bayesian Network with 3 time intervals.

A. **Bayesian Network**

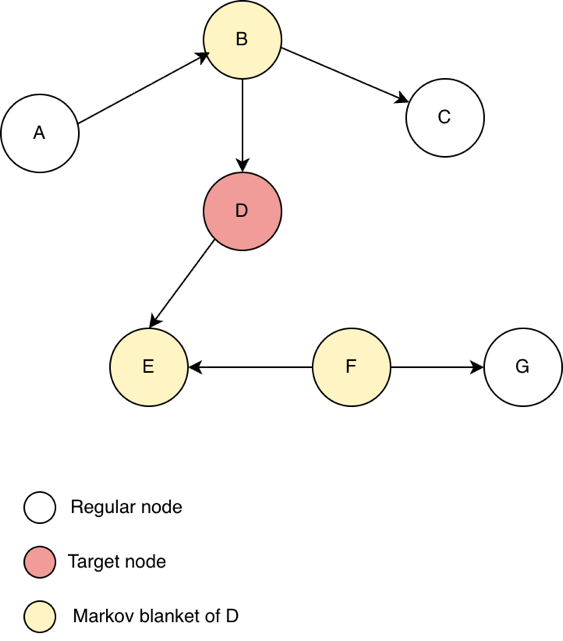

B. **Dynamic Bayesian Network**

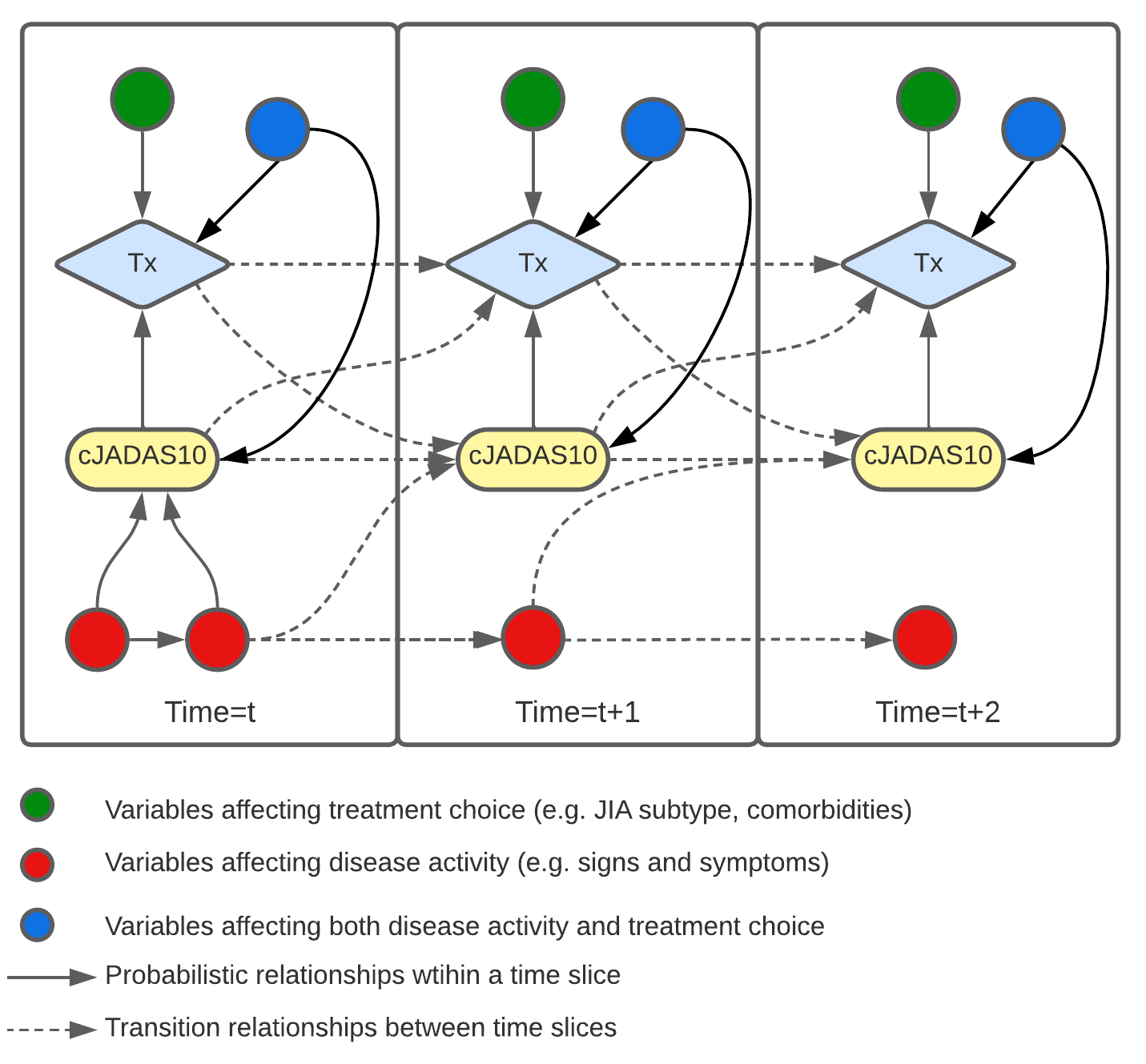
