## Supplementary Figure 2 for "Machine Learning Prediction of Disease Trajectories for Children with Juvenile Idiopathic Arthritis"

**Supplementary Figure 2.** **Learning curves of a feedforward neural net for prediction of remission at 12 months.** As the neural network’s weights were iteratively updated, training set accuracy improved, whereas performance on unseen datasets began to deteriorate after the early training phase, consistent with model overfitting.

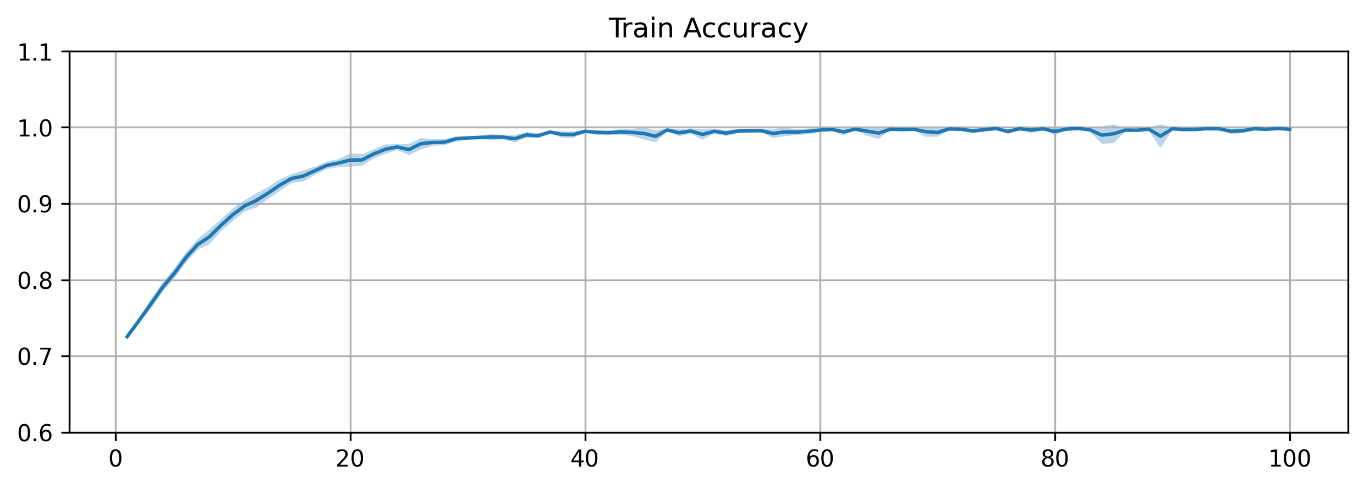

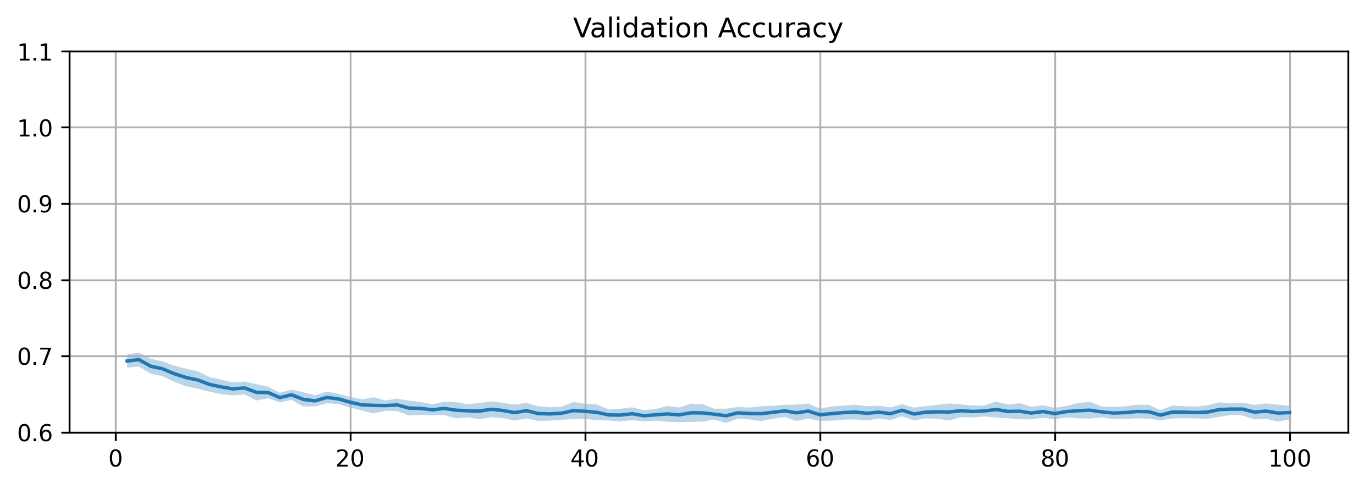

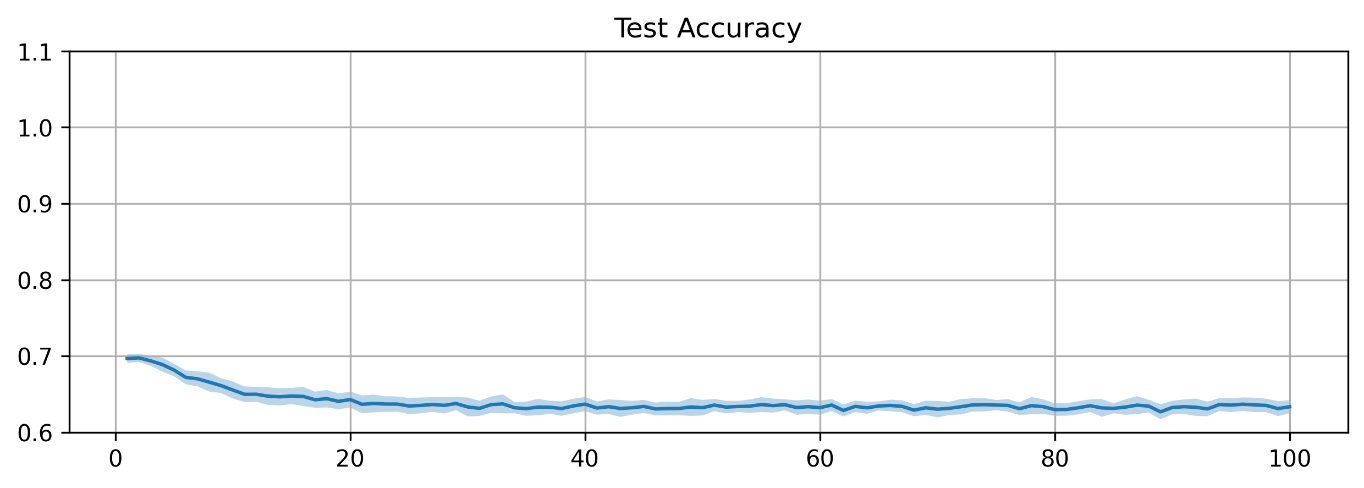
