## Supplementary Table 1 for "Machine Learning Prediction of Disease Trajectories for Children with Juvenile Idiopathic Arthritis"

**Supplementary Table 1. Variables used for model development**

| **Variables** | **Representation in the CARRA Registry** | **Categorical representation in DBN** |
| --- | --- | --- |
| JIA category | Categorical | Enthesitis-related arthritis, Oligoarthritis, polyarthritis (RF -ve), polyarthritis (RF +ve), psoriatic arthritis, undifferentiated arthritis |
| Gender | Categorical | Female, male |
| Self-reported race | Categorical | American Indian, Asian, Black, Middle Eastern, Native Hawaiian, White |
| Self-reported ethnicity | Categorical | Hispanic, non-Hispanic |
| Household income | Categorical | <25000, 25-49999, 50-74999, 75-99999, 100000-150000, >150000, prefer not to answer |
| Highest level of parent/guardian education completed | Categorical | Elementary/middle school, some high school, graduated high school, college or technical school, graduate school, masters/doctorate or professional degree, prefer not to answer |
| Insurance | Categorical | Medicare, Medicaid, private, uninsured, non-US |
| Time from symptom onset to diagnosis (months)* | Continuous | ≤2, >2 to 5, >5 to 12, >12 |
| Age at JIA Diagnosis | Continuous | ≤6, >6 to ≤12, >12 |
| cJADAS10[[7]](https://sciwheel.com/work/citation?ids=10940379&pre=&suf=&sa=0&dbf=0) | Continuous | Polyarticular JIA (≤2.5 inactive; >2.5-5 minimal; >5-16 moderate; and >16 high disease activity)  Oligoarticular JIA (≤1.1 inactive disease; >1.1-4 minimal; >4-12 moderate; and >12 high disease activity |
| bDMARD | Categorical | Yes, no |
| csDMARD | Categorical | Yes, no |
| Glucocorticoids | Categorical | Yes, no |
| HLAB27 | Categorical | Yes, no |
| Trisomy21 | Categorical | Yes, no |
| Family history of rheumatic diseases | Categorical | Yes, no |
| Family history of inflammatory bowel disease | Categorical | Yes, no |
| Family history of uveitis | Categorical | Yes, no |
| Family history of other autoimmune disorder | Categorical | Yes, no |
| Anti-CPP | Categorical | Yes, no |
| ANA | Categorical | Yes, no |
| Total active joint | Categorical | None, >5, >=5 |
| Morning stiffness | Categorical | ≤15 minutes, 16-60 minutes, >60 minutes, None |
| Swelling | Categorical | Yes, no |
| Joint stiffness | Categorical | Yes, no |
| Active enthesitis | Categorical | Yes, no |
| Active sacroiliitis | Categorical | Yes, no |
| Schober test | Categorical |  |
| Radiographic joint damage | Categorical | Yes, no |
| Sacroiliac joint damage | Categorical | Yes, no |
| Sacroiliac arthritis | Categorical | Yes, no |
| Uveitis | Categorical | Yes, no |
| Psoriasis | Categorical | Yes, no |
| Sacroiliac tenderness | Categorical | Yes, no |
| Inflammatory back pain | Categorical | Yes, no |
| Abnormal ESR | Categorical | Yes, no |
| Abnormal CRP | Categorical | Yes, no |
| Primary immunodeficiency | Categorical | Yes, no |
| Asthma | Categorical | Yes, no |
| Other autoimmune disorder | Categorical | Yes, no |

*****Grouped into four categories with equal numbers of observations.
