## Supplementary Table 2 for "Machine Learning Prediction of Disease Trajectories for Children with Juvenile Idiopathic Arthritis"

**Supplementary Table 2: Medications**

| **Categories** | **Medications** |
| --- | --- |
| csDMARD | Methotrexate, leflunomide, hydroxychloroquine, sulfasalazine |
| b/tsDMARD |  |
| Anti-IL1 | Anakinra, canakinumab, rilonacept |
| Anti-IL6 | Tocilizumab |
| TNFi | Adalimumab, certolizumab, etanercept, golimumab, infliximab |
| Others | Ixekizumab, secukinumab, vedolizumab, guselkumab, risankizumab |
| Small molecule inhibitors | Apremilast, tofacitinib |
| Glucocorticoids | Prednisolone, prednisone, methylprednisolone, dexamethasone |
