## Supplementary Table 3 for "Machine Learning Prediction of Disease Trajectories for Children with Juvenile Idiopathic Arthritis"

**Supplementary Table 3. Hyperparameters of Deep Learning**

**Fully convolutional neural network**

Training-related hyperparameters

- Maximum training epochs: 300
- Size of mini batch: 32
- Learning rate: 0.00025
- L1 regularization scale λ: 0.001
- Weight initialization method: He initialization

Architecture-related hyperparameters

- Number of layers: 4 convolutional layers
- Number of filters (channels) from input to output layer: 16, 32, 8, 2
- Sizes of kernels from input to output layer: 7x2, 1x1, 1x1, 1x1
- Stride step size: 1
- Zero padding: “Same” output size
- Pooling: No pooling
- Activations except output layer: ReLU

**Dual-branch Network**

Training-related hyperparameters

- Maximum training epoch: 1000
- Size of mini batch: 32
- Learning rate: 0.00025
- L1 regularization scale λ: 0.0001
- Weight initialization method: He initialization

Architecture-related hyperparameters

- Number of layers in medication feature extractor: 2 fully connected layers
- Number of hidden units in medication feature extractor: 32, 32
- Number of layers in status feature extractor: 2 fully connected layers
- Number of hidden units in status feature extractor: 32, 32
- Number of layers in fusion network: 2 fully connected layers
- Number of hidden units in fusion network: 16, 2
- Activations except output layer: ReLU

**Feedforward Network**

Training-related hyperparameters

- Maximum training epoch: 1000
- Size of mini batch: 32
- Learning rate: 0.00025
- L1 regularization scale λ: 0.00001
- Weight initialization method: He initialization

Architecture-related hyperparameters

- Number of layers: 4 fully connected layers
- Number of hidden units: 64, 32, 16, 2
- Activations except output layer: ReLU
