## Supplementary Table 4 for "Machine Learning Prediction of Disease Trajectories for Children with Juvenile Idiopathic Arthritis"

**Supplementary Table 4.** **Comparison between neural network architectures.** Accuracy and AUC on the test dataset for four neural network architectures predicting remission at 12 months. The different architectures achieved comparable performance.

| Metrics | Feedforward neural net | Double-branch with  additive interaction | Double-branch with  multiplicative interaction | Double-branch net with  concatenation |
| --- | --- | --- | --- | --- |
| Accuracy | 0.70 | 0.70 | 0.70 | 0.71 |
| AUC | 0.76 | 0.77 | 0.77 | 0.77 |
