## Supplementary Table 5 for "Machine Learning Prediction of Disease Trajectories for Children with Juvenile Idiopathic Arthritis"

**Supplementary Table 5. Patterns of missing cJADAS10**

| **Month** | **Total enrolled, n** | **% missing cJADAS10** |
| --- | --- | --- |
| 0 | 11889 | 23.3 |
| 6 | 10683 | 38.1 |
| 12 | 10297 | 40.9 |
| 18 | 9890 | 46.7 |
| 24 | 9521 | 50.8 |
| 30 | 9063 | 55.2 |
| 36 | 8658 | 57.8 |
