## Supplementary Methods for "Machine Learning Prediction of Disease Trajectories for Children with Juvenile Idiopathic Arthritis"

**Supplementary Methods: Model Development**

*Dynamic Bayesian Network*

DBN learning employed a two-phase approach, combining structure discovery with probabilistic parameter estimation. In the structure learning phase, we implemented a Tabu search algorithm to identify the optimal network topology by systematically evaluating candidate structures through edge addition, removal, and reversal operations.[^1^](https://sciwheel.com/work/citation?ids=9114855&pre=&suf=&sa=0&dbf=0) The algorithm maintains a tabu list of recently explored configurations to prevent local optima cycling, while scoring each candidate structure using the Bayesian Information Criterion (BIC) to balance model fit against complexity. Temporal constraints were incorporated to ensure plausible relationships among variables. To enable learning from incomplete data, we applied Bayesian parameter estimation using the Expectation-Maximization (EM) algorithm to learn the conditional probability distributions governing variable relationships following structural learning.[^2^](https://sciwheel.com/work/citation?ids=6462061&pre=&suf=&sa=0&dbf=0) The EM approach alternates between estimating hidden disease states (i.e. aspects of a disease that we cannot directly observe in the data but that still affect what we do observe) given current parameters (E-step) and updating parameters to maximize data likelihood given estimated states (M-step), iterating until convergence. This methodology yields transition probabilities governing temporal disease state evolution and conditional probability tables capturing complex variable dependencies, all while maintaining interpretability essential for clinical decision support applications.

*Convolutional Neural Network (CNN)*

We investigated multiple CNN architectures to identify the optimal model configuration. Our exploration included: (1) a standard feedforward neural network, (2) a fully convolutional network that eliminates fully connected layers, and (3) a dual-branch architecture with separate input pathways. The dual-branch network consists of two parallel feature extraction sub-networks, one dedicated to medication-related features and the other to non-medication features, followed by a fusion sub-network that combines these representations to generate predictions. This architecture allows the model to learn specialized representations for each feature type, capturing distinct patterns that might be lost if all input were processed jointly.[^3,4^](https://sciwheel.com/work/citation?ids=6733701,18365246&pre=&pre=&suf=&suf=&sa=0,0&dbf=0&dbf=0) To merge the medication and non-medication feature representations, we evaluated three fusion strategies: additive interaction (element-wise addition), multiplicative interaction (element-wise multiplication), and concatenation (joining features along the channel dimension). This systematic comparison allowed us to assess how different architectural choices and feature fusion methods impact model performance. In each experimental setup, we optimized network parameters using the Adam (Adaptive Moment Estimation) algorithm. We used 12-month outcome prediction as our benchmark task for comparing different architectural configurations. Once the optimal architecture was identified through this evaluation, we extended the final model to predict outcomes at all available time points.

We first developed CNN models using the full set of variables (see Supplementary Table 1). To mitigate potential overfitting, we subsequently developed models using a subset of features identified to be predictive by the DBN models. In all analyses, continuous variables were log-transformed to reduce skewness, and categorical variables were encoded using one-hot encoding. Supplementary Table 3 summarizes the hyperparameters used in model development.

To further mitigate overfitting, we also adopted two regularization techniques: L1 regularization and dropout.[^5,6^](https://sciwheel.com/work/citation?ids=16679086,11747145&pre=&pre=&suf=&suf=&sa=0,0&dbf=0&dbf=0) To implement L1 regularization, we augmented the standard cross-entropy loss function with an L1 penalty term that penalizes the absolute values of network weights. This modification serves dual purposes: (1) it constrains model complexity by encouraging sparse weight distributions, thereby reducing overfitting, and (2) it promotes model sparsity by driving many weights to exactly zero, which can aid interpretation by identifying which network connections are essential for prediction. The regularization strength was controlled by a hyperparameter λ, which we tuned during model selection. To implement dropout, we randomly deactivated 50% of neurons and their connections at each forward pass. This stochastic perturbation prevents neurons from developing complex interdependencies that only work on training data, thereby reducing overfitting. 50% dropout rate has been shown to be optimal for a wide range of networks and tasks.[^6^](https://sciwheel.com/work/citation?ids=11747145&pre=&suf=&sa=0&dbf=0)

*Sensitivity analyses*

To improve predictive performance of our models, we conducted a number of sensitivity analyses, including: (a) confining the study cohort to those enrolled within the first 6 months of diagnosis to create an inception cohort; (b) confining the study cohort to those enrolled for at least 2 or 3 years to ensure adequate follow-up; (c) developing separate models for patients with polyarticular course and those with oligoarticular course JIA; (d) confining the study cohort to those enrolled in the STOP-JIA (Start Time Optimization of Biologics in Polyarticular JIA) study, whereby patients were prospectively followed on one of three consensus treatment plans to compare strategies for initiating biologics; and (e) developing separate models for predicting outcome at each time point (12, 18, 24, 30, 36 months from registry enrollment), instead of a joint model; (f) expressing medication use as cumulative days of exposure, instead of as binary variables; and (g) categorizing medications in finer categories, including tumor necrosis factor inhibitors (TNFi), anti-interleukin(IL)6, other bDMARDs, methotrexate, other csDMARDs and tsDMARDs. Additionally, we explored several approaches to address missing data, including zero imputation, autoencoder, MissForest, DataWig, MICE (Multivariate Imputation by Chained Equations) and MIDAS (Multiple Imputation with Denoising Autoencoders).[^7–12^](https://sciwheel.com/work/citation?ids=16816521,18360542,3014571,18360546,10617501,18360565&pre=&pre=&pre=&pre=&pre=&pre=&suf=&suf=&suf=&suf=&suf=&suf=&sa=0,0,0,0,0,0&dbf=0&dbf=0&dbf=0&dbf=0&dbf=0&dbf=0)

**Supplementary References**

[1     Friedman N, Nachman I, Pe’er D. Learning Bayesian network structure from massive datasets: The" sparse candidate" algorithm. *arXiv preprint arXiv:13016696* 2013.](https://sciwheel.com/work/bibliography/9114855)

[2     Dempster AP, Laird NM, Rubin DB. Maximum Likelihood from incomplete data Via the *EM* algorithm. *Journal of the Royal Statistical Society: Series B (Methodological)* 1977; **39**: 1–22.](https://sciwheel.com/work/bibliography/6462061)

[3     Feichtenhofer C, Pinz A, Zisserman A. Convolutional Two-Stream Network Fusion for Video Action Recognition. In: 2016 IEEE Conference on Computer Vision and Pattern Recognition (CVPR). IEEE, 2016: 1933–41.](https://sciwheel.com/work/bibliography/6733701)

[4     Wang Y, Song J, Wang L, Gool L, Hilliges O. Two-Stream SR-CNNs for Action Recognition in Videos. In: Procedings of the British Machine Vision Conference 2016. British Machine Vision Association, 2016: 108.1-108.12.](https://sciwheel.com/work/bibliography/18365246)

[5     Moradi R, Berangi R, Minaei B. A survey of regularization strategies for deep models. *Artif Intell Rev* 2020; **53**: 3947–86.](https://sciwheel.com/work/bibliography/16679086)

[6     Srivastava N, Hinton G, Krizhevsky A. Dropout: a simple way to prevent neural networks from overfitting. *The journal of machine* 2014.](https://sciwheel.com/work/bibliography/11747145)

[7     Cardoso Pereira R, Seoane Santos M, Pereira Rodrigues P, Henriques Abreu P. Reviewing autoencoders for missing data imputation: technical trends, applications and outcomes. *jair* 2020; **69**: 1255–85.](https://sciwheel.com/work/bibliography/16816521)

[8     Jin H, Jung S, Won S. missForest with feature selection using binary particle swarm optimization improves the imputation accuracy of continuous data. *Genes Genomics* 2022; **44**: 651–8.](https://sciwheel.com/work/bibliography/18360542)

[9     Stekhoven DJ, Bühlmann P. MissForest — non-parametric missing value imputation for mixed-type data. *Bioinformatics* 2012; **28**: 112–8.](https://sciwheel.com/work/bibliography/3014571)

[10    Zaninotto P. Multiple imputation by chained equations (MICE). Instats Inc., 2024 DOI:10.61700/1TR36KP5GWA5B1858.](https://sciwheel.com/work/bibliography/18360546)

[11    Lall R, Robinson T. The MIDAS Touch: Accurate and Scalable Missing-Data Imputation with Deep Learning. *Political Analysis* 2021; : 1–18.](https://sciwheel.com/work/bibliography/10617501)

[12    Biessmann F, Rukat T, Schmidt P, *et al.* DataWig: Missing Value Imputation for Tables. *J Mach Learn Res* 2019; **20**: 1–6.](https://sciwheel.com/work/bibliography/18360565)

[13    Trincianti C, van Dijkhuize EHP, Alongi A, *et al.* Definition and validation of the American College of Rheumatology 2021 JADAS cutoffs for Disease Activity States in Juvenile Idiopathic Arthritis. *Arthritis Rheumatol In press* 2021.](https://sciwheel.com/work/bibliography/10940379)
